# Epigenetic Regulation of Immune Dysfunction in Chronic Prostatitis/Chronic Pelvic Pain Syndrome (CP/CPPS)

**DOI:** 10.64898/2026.01.14.26344138

**Authors:** Praveen Thumbikat, Goutham Pattabiraman, Farzaneh Sharifzad, Yongyong Yang, Zhiqiang Liu, Catherine V Osborn, Stephen F Murphy, Qi Cao, Anthony J. Schaeffer

**Author notes:** Corresponding Author: Praveen Thumbikat, D.V.M, Ph.D., Professor, Department of Urology, Feinberg School of Medicine, Searle 12-562, 320 E. Superior St., Chicago, IL 60611. **AUTHOR CONTRIBUTIONS:** Praveen Thumbikat-directed study, analyzed data, prepared manuscript Goutham Pattabiraman – performed study and analyzed data. Farzaneh Sharifzad, Catherine Osborn Stephen Murphy, Yongyong Yang and Zhiqiang Liu - performed study and analyzed data. Qi Cao and Anthony Schaeffer – performed study, reviewed manuscript. DATA AVAILABILITY: Data developed during this study are available on request. Clinical data made available, upon request, will follow established protocol guidance by Northwestern University IRB. PATIENT CONSENT and ETHICS APPROVAL: The Northwestern University Institutional Review Board (Panel D) approved the protocols (IRB STU00030121, STU00202831 and STU 00215831), and all participants provided written informed consent. All animal experiments procedures were approved by the Northwestern University Animal Care and Use Committee. Conflicts of interest:* The authors have no conflicts of interest to declare relevant to the data or results presented in this manuscript.

## Abstract

**Background:** Chronic prostatitis/chronic pelvic pain syndrome (CP/CPPS) is a prevalent and debilitating condition with unclear etiology. Increasing evidence implicates immune dysregulation, yet the molecular mechanisms underlying impaired immune regulation remain poorly defined. This study investigated the role of adaptive immune responses and DNA methylation in CP/CPPS pathogenesis.

**Methods:** Voided bladder 3 (VB3) urine samples from CP/CPPS patients and healthy controls were analyzed for CD4⁺ T cell markers and lineage-defining transcription factors. DNA methylation profiling of peripheral blood mononuclear cells (PBMCs) and purified CD4⁺ T cells was performed using targeted methylation arrays. Functional assays evaluated IL-10 production following lipopolysaccharide (LPS) stimulation, with or without azacitidine (AZA), a DNA methyltransferase inhibitor that reverses methylation-dependent gene silencing. In vivo relevance was assessed using the experimental autoimmune prostatitis (EAP) mouse model.

**Results:** VB3 samples from CP/CPPS patients demonstrated elevated CD4⁺ T cell transcripts with a Th17/Th1 (RORγT⁺/TBET⁺) bias and reduced FOXP3 expression. DNA methylation analysis revealed hypermethylation of IL10 and FOXP3 promoters and hypomethylation of inflammatory genes including CD274, ITGAL, and TNFα. PBMCs from patients exhibited diminished IL-10 secretion in response to LPS, which was restored by azacitidine treatment. In the EAP model, recombinant IL-10 administration failed to attenuate pelvic allodynia, whereas azacitidine significantly reduced pain sensitivity.

**Conclusions:** CP/CPPS is associated with widespread epigenetic reprogramming of immune regulatory genes leading to impaired IL-10–mediated immune regulation. Pharmacologic inhibition of DNA methylation restored IL-10 responses and alleviated pain in vivo, supporting demethylation therapy as a potential strategy for treating chronic prostatic inflammation and pelvic pain.

## Introduction

Chronic prostatitis/chronic pelvic pain syndrome (CP/CPPS) is a common condition within the spectrum of urologic chronic pelvic pain syndrome (UCPPS), diagnosed exclusively in adult men [1; 8; 29]. Despite its high prevalence and impact on quality of life, the etiology and pathogenesis of CP/CPPS remain poorly understood. The condition is typically diagnosed by exclusion and is characterized by persistent pelvic pain, often accompanied by lower urinary tract symptoms and sexual dysfunction.

Over the past two decades, multiple hypotheses have been proposed to explain CP/CPPS pathophysiology, including autoimmunity, hormonal imbalances, and pelvic floor dysfunction [21]. More recently, neuroimaging studies have identified altered neuronal density and connectivity in the brains of CP/CPPS patients, although it remains unclear whether these are primary drivers or secondary adaptations to pathology elsewhere in the body [35].

Experimental studies in animal models have demonstrated that immune responses directed at the prostate are sufficient to induce hallmark symptoms of pelvic pain and voiding dysfunction [7; 17; 19; 20; 26; 28]. While the precise immune triggers vary—some involving IFN-γ–driven pathways [18], others, including work from our group, implicating IL-17–mediated responses [4; 17; 18; 20] —a unifying feature across models is the activation of adaptive immunity, particularly T cell-mediated inflammation.

In chronic immune-mediated diseases, a growing body of evidence highlights the role of epigenetic modifications in shaping immune susceptibility. Epigenetic mechanisms, including chromatin remodeling, non-coding RNAs, and DNA methylation, control gene expression without altering the underlying DNA sequence and are responsive to developmental, metabolic, and environmental inputs [11; 22]. Among these, DNA methylation is the most extensively studied and involves covalent modification of cytosine residues within CpG islands, catalyzed by DNA methyltransferases (DNMTs) and interpreted by methyl-CpG-binding proteins [16] [3; 5; 31].

Aberrant DNA methylation patterns have been implicated in a wide array of pathological states by disrupting immune tolerance or promoting inflammatory gene expression [27]. Epigenome-wide association studies (EWAS) have linked methylation changes to environmental exposures such as chemical toxins and smoking, as well as to diseases characterized by chronic inflammation, including inflammatory bowel disease, Alzheimer’s disease, cancer, and diabetes [25] [13] [34] [6]. Across chronic pain states, DNA methylation emerges as a key, durable regulator of gene programs that stabilize pain long after the initiating insult has resolved [37]. Peripheral nerve injury and other persistent pain models show coordinated methylation shifts at promoters controlling neurotransmission, intracellular signaling, and immune pathways, with methylation often inversely tracking mRNA expression and correlating with pain intensity [15]. Critically, T-cell methylation signatures substantially overlap with pain-linked patterns in prefrontal cortex, and compact T-cell methylation panels can classify neuropathic pain and predict mechanical hypersensitivity [15]. These findings position peripheral immune cells as minimally invasive windows into central pain biology while highlighting methylation-encoded states as actionable targets for disease modification [15; 37].

Given these observations, we hypothesized that aberrant DNA methylation contributes to the immune imbalance observed in CPPS. We further posited that these changes disproportionately affect regulatory immune pathways. This is based on the observation that multiple distinct proinflammatory mechanisms—including IL-17 and IFN-γ—can induce pelvic pain in animal models, suggesting that failure of regulatory restraint, rather than the nature of the initial immune stimulus, may be a common determinant of chronic disease persistence.

## Methods

### Study population and specimen collection

Men ≥ 18 years old who were healthy or had a prior diagnosis of Category III chronic prostatitis/chronic pelvic pain syndrome (CP/CPPS) recruited through the Northwestern University Feinberg School of Medicine Department of Urology via physician referral, advertisements, and ClinicalTrials.gov outreach (NCT01676857, NCT03167216, NCT05185180). Eligibility for CP/CPPS patients included the presence of pelvic pain or discomfort for ≥ 3 months within the preceding 6 months and a National Institutes of Health-Chronic Prostatitis Symptom Index (NIH-CPSI) total score ≥ 12/43 at screening. Dr A. Schaeffer and colleagues collected questionnaire data, Voided Bladder 3 (VB3) urine, and blood samples at the Northwestern Memorial Hospital outpatient urology clinic, Chicago, IL. The Northwestern University Institutional Review Board (Panel D) approved the protocols (IRB STU00030121, STU00202831 and STU 00215831), and all participants provided written informed consent.

VB3 specimens were centrifuged (300 g, 4 min, 4 °C); supernatants were stored at −80 °C, and cell pellets were resuspended and used for RNA extraction. Peripheral blood was drawn into BD Vacutainer® CPT™ tubes pre-loaded with Ficoll and processed within 2 h by centrifugation (1,800 × g, 20 min, room temperature), to isolate the peripheral-blood mononuclear cell (PBMC) layer, which was cryopreserved till further use, in liquid nitrogen. Storage and handling conditions were standardized between controls and CPPS patients.

### Quantitative Reverse-Transcription PCR (qRT-PCR)

Total RNA was extracted from the VB3 cells with TRIzol reagent (Invitrogen) and reverse-transcribed with qScript™ cDNA SuperMix (QuantaBio) following the manufacturers’ protocols. qRT-PCR reactions were prepared with PerfeCTa® SYBR Green SuperMix (QuantaBio) and run on a CFX Connect™ Real-Time PCR System (Bio-Rad). Primer pairs were designed in silico using NCBI Primer-BLAST. Relative transcript levels were calculated by the 2^-ΔΔCt method, with subsequent data visualization and statistical analysis performed in GraphPad Prism (GraphPad Software).

### CD4 T cell extraction and DNA methylation studies

The PBMC band was thawed, washed, assayed for adequate cell viability and used for enrichment of CD4⁺ T cells using the EasySep™ Human CD4⁺ T Cell Isolation Kit (STEMCELL Technologies) following the manufacturer’s negative-selection protocol. Purified cells were then pelleted and shipped on dry ice to EpigenDx (Hopkinton, MA). EpigenDx performed bisulfite conversion and targeted Next-Gen Bisulfite Sequencing followed by locus-specific PCR (NGS070V3 - Human FOXP3 Regulators and additional genes) and pyrosequencing to quantify CpG methylation according to their standard validated protocols.

### Cell culture studies

PBMCs were thawed in a 37°C water bath and slowly diluted with pre-warmed RPMI 1640 medium supplemented with 10% fetal bovine serum. After centrifugation at 300 × g for 5 minutes, the cells were resuspended in fresh culture medium and incubated at 37°C with 5% CO₂ for one hour to allow recovery. For *in. vitro* inflammation studies, 0.5× 10^5^ cells/well in 6-well plates were stimulated with 1 μg/mL /well of LPS (Sigma); and incubated for 24 hours at 37°C, 5% CO₂. Supernatants were collected by centrifugation at 300 × g for 5 minutes and stored at −80°C. IL-10 levels were quantified using the Quantikine Human IL-10 ELISA Kit (R&D Systems, Catalog #D1000B) according to the manufacturer’s instructions. For *in vitro* methylation inhibition studies, PBMC’s were treated with 1uM DNA methyltransferase inhibitor, AZA (azacitidine) for 48 or 96 hours followed by isolation of total RNA and *qRT-PCR* for IL10 expression.

### Therapeutic studies in experimental autoimmune prostatitis (EAP)

CP/CPPS was modeled using mice with experimental autoimmune prostatitis (EAP) as previously described [28]. Animal experiments procedures were approved by the Northwestern University Animal Care and Use Committee. Briefly, male NOD/ShiLtJ mice (5–7 weeks old) were immunized subcutaneously with 100μg of rat prostate antigen emulsified in TiterMax Gold adjuvant (1:1 ratio, GA 30093) (day 0) and allowed to develop autoimmune mediated prostate inflammation over 28 days. Behavioral assessment of changes in mechanical allodynia of the pelvic region were quantified from von Frey filament testing on days 0, 7, 14, 21, and 28 post-immunization. To evaluate therapeutic effects of IL10, day 22 to day 27, mice received daily intraperitoneal injections of 1μg recombinant mouse IL-10 in 100μL PBS while controls received vehicle injections (sterile PBS). For AZA therapeutic assessment, male C57BL/6J mice (5-7) weeks of age were allowed to develop EAP and treatment was initiated at 28 days after EAP by intraperitoneal injection of 5-AZA (A3666, Sigma) at 2.5mg/kg. Injections were repeated daily for 7 days. Behavioral assessment of changes in mechanical allodynia of the pelvic region were quantified from von Frey filament testing at the onset of EAP, and at the beginning and end of the treatment regimen.

### Statistical analyses

Statistical analyses were performed using GraphPad Prism™ (GraphPad Software, San Diego, CA, USA). Statistical tests utilized in each experiment, are indicated in figure legends (in most figures, each dot represents individual patients/animals). Data are represented as the mean ± standard deviation (SD) or mean ± standard error of the mean (S.E.M.) as appropriate. * p < 0.05, ** p < 0.01, *** p <0.001, **** p < 0.0001.

## RESULTS

### Altered adaptive immunity in CP/CPPS

To assess local adaptive immune responses, we profiled CD4⁺ T-cell–associated transcripts in voided bladder 3 (VB3) urine sediments from men with chronic prostatitis/chronic pelvic pain syndrome (CP/CPPS; *n* = 24) and healthy controls (*n* = 4; NU IRB STU00030121). While the limited numbers of immune cells in VB3 sediment precluded analytical methods such as flow cytometry, quantitative RT–PCR revealed a significant increase in CD4 mRNA expression in CP/CPPS samples compared with controls (*p* = 0.0154; *Fig. 1a*), consistent with either enhanced local CD4⁺ T-cell recruitment or CD4⁺ T-cell activation. To examine T helper subset polarization, we studied expression of lineage-defining transcription factors—TBET (Th1), RORγT (Th17), GATA3 (Th2), and FOXP3 (Treg)—normalized to CD4 expression. Among these, RORγT was significantly upregulated in patients (*p* = 0.0041; *Fig. 1b–e*), whereas TBET, GATA3, and FOXP3 showed no consistent group-level differences. Notably, expression profiles displayed marked inter-individual heterogeneity. Z-score analysis revealed distinct patient subsets with elevated RORγT and GATA3 and reduced FOXP3 and TBET expression relative to the control mean (*Fig. 1f*). Collectively, these data indicate that prostate-localized immune activity in CP/CPPS is characterized by a Th17/Th2-biased and Treg/Th1-deficient profile, suggesting an imbalance in adaptive immune regulation that may contribute to the chronic inflammatory milieu underlying pelvic pain.

**Figure 1.**
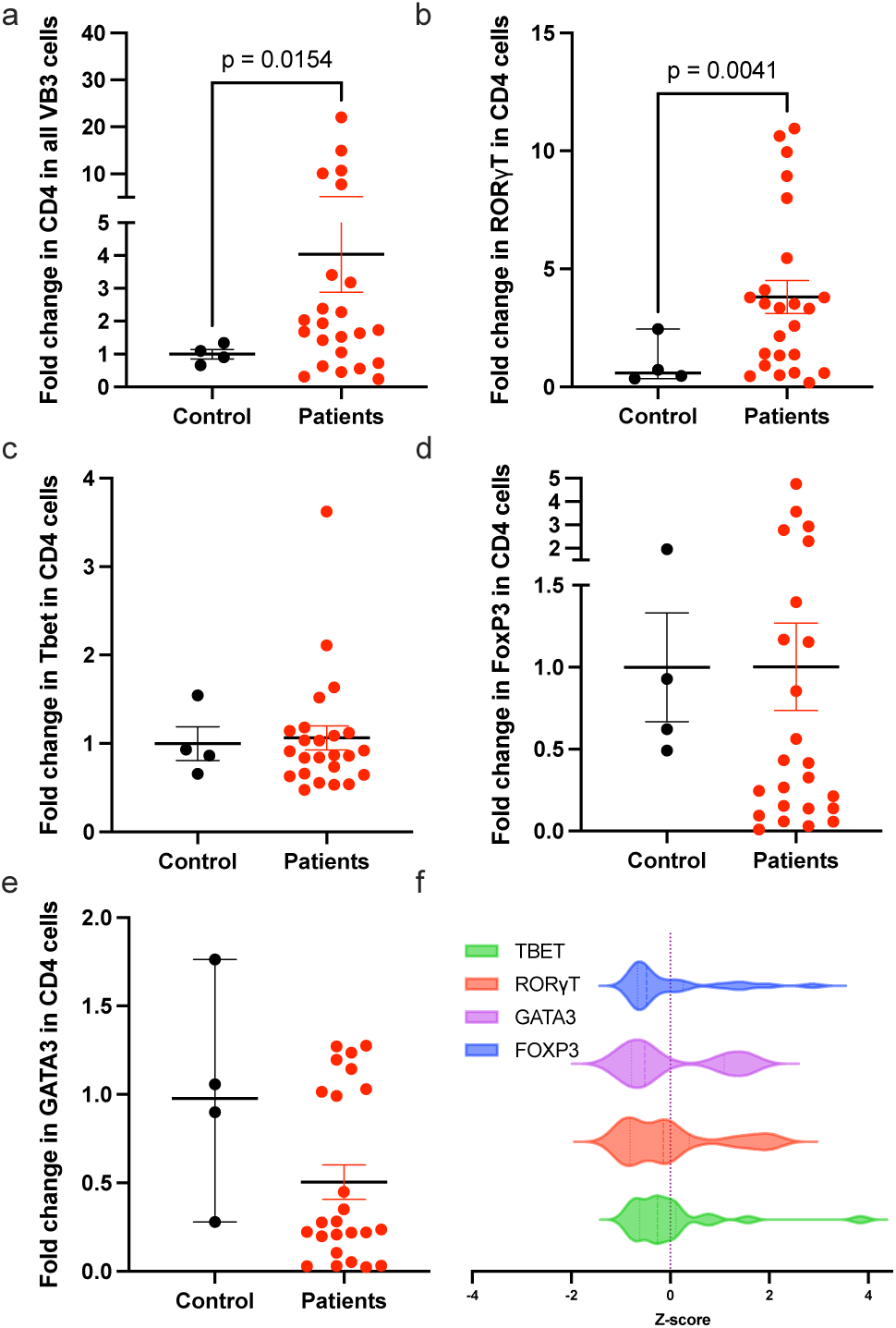
Prostate-localized CD4 T cell subsets demonstrate altered immune responses. Cells were isolated from voided bladder 3 (VB3) urine samples of CP/CPPS patients (n = 24) and healthy controls (n = 4) and analyzed by qRT-PCR for genes associated with adaptive immune function. (a) CD4 mRNA expression; (b–e) expression of key transcription factors: TBET (Th1), RORγT (Th17), GATA3 (Th2), and FOXP3 (T-reg), each normalized to CD4 expression. Statistical comparisons were performed using two-tailed unpaired t-tests; individual p-values are shown. (f) Z-scores were calculated as *(X − mean) / standard deviation*, where a Z-score of 0 indicates a value equivalent to the control mean, and positive or negative scores reflect the magnitude of deviation.

### Aberrant DNA methylation signatures in PBMCs from CP/CPPS patients

To determine whether systemic immune dysregulation in CP/CPPS is associated with epigenetic alterations, we analyzed peripheral blood mononuclear cells (PBMCs) from CP/CPPS patients (n = 10) and age-matched healthy controls (n = 10) using a targeted methylation array focused on regulatory immune genes. Multiple loci displayed significant differences in DNA methylation between patients and controls. The IL10 promoter was hypermethylated at three CpG islands (#4, 3 and 2.1) located at –387, –385, and –355 from the ATG start site (Fig. 2a). This is consistent with reduced expression of this anti-inflammatory cytokine observed in previous studies. Additional hypermethylation was detected in PDL2 and MYC (Fig. 2b, c), whereas CD274 (PD-L1), ITGAL (LFA-1), and TNFA loci were hypomethylated (Fig. 2d–f). Although the absolute methylation differences at individual sites were modest, the overall pattern indicated coordinated remodeling of immune-regulatory genes in PBMCs from CP/CPPS patients. These data support the presence of systemic epigenetic alterations that may contribute to the proinflammatory immune phenotype reported in CPPS cohorts.

**Figure 2.**
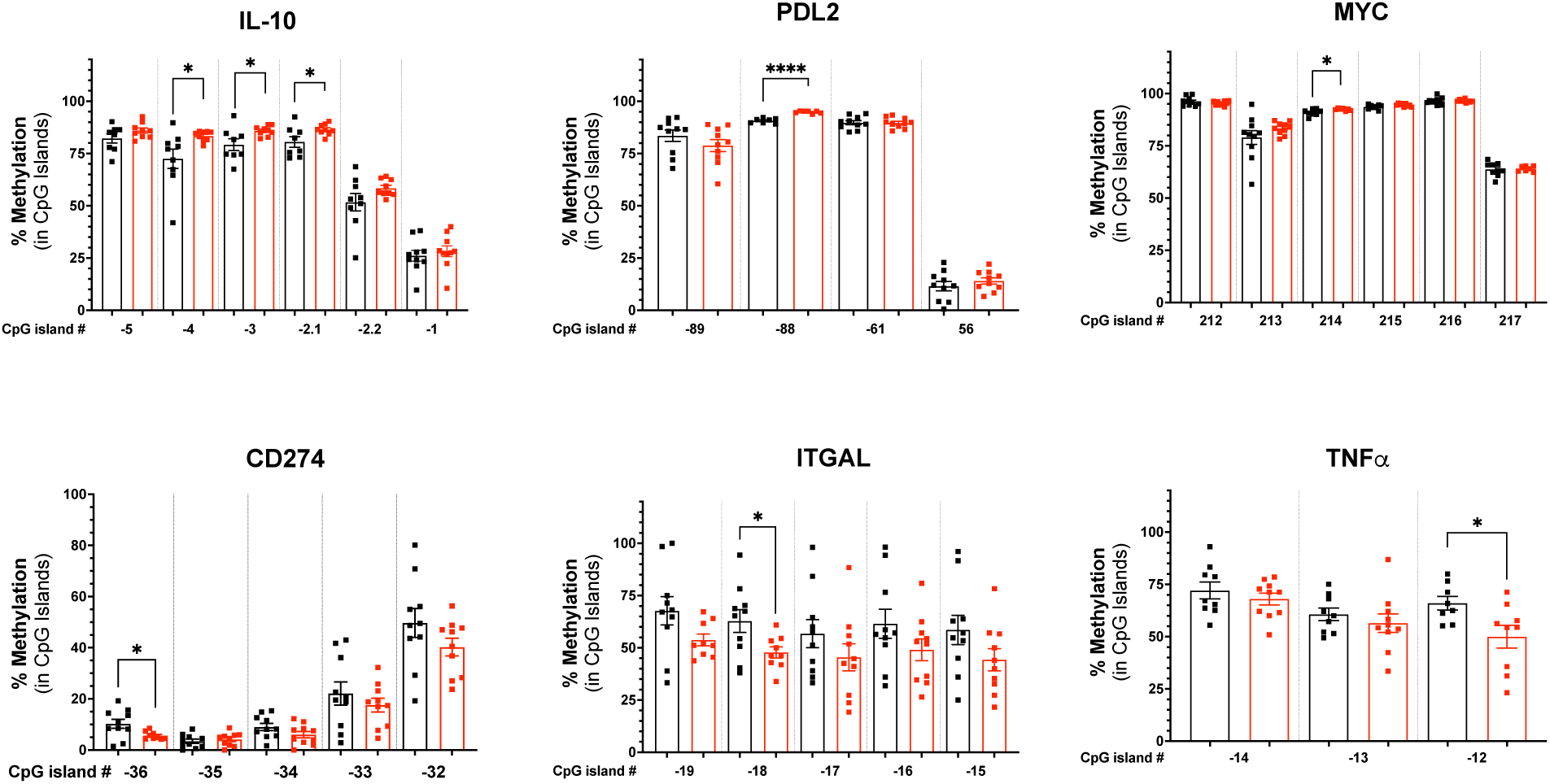
Aberrant DNA methylation signatures in PBMCs from CP/CPPS patients. Peripheral blood mononuclear cells (PBMCs) from a separate cohort of CP/CPPS patients (n = 10) and healthy controls (n = 10) were analyzed for DNA methylation changes in genes linked to FOXP3 expression and T-regulatory function using the EpigenDx Human FOXP3 array. Methylation at multiple CpG sites was assessed and compared between groups. Statistical comparisons between groups were performed using two-tailed unpaired t-tests; *p<0.05, **p<0.01, ***p<0.001, ****p<0.0001.

### Expanded methylation changes in purified CD4⁺ T cells

Given the central role of CD4⁺ T cells in models of CP/CPPS pathogenesis [26; 28], we next examined whether the DNA methylation alterations observed in PBMCs were associated with this subset. Purified CD4⁺ T cells from CP/CPPS patients (n = 9) and age-matched healthy controls (n = 8) were analyzed using a targeted methylation array focused on regulatory immune loci. Consistent with the PBMC findings, IL10 promoter hypermethylation was observed in CD4⁺ T cells from CP/CPPS patients (Fig. 3). In addition, FOXP3 promoter hypermethylation was detected at the CNS1 and CNS2 regions, which are critical for maintaining regulatory T cell stability. These FOXP3 changes were not evident in bulk PBMCs, indicating that cell-specific analysis reveals regulatory lesions masked in mixed populations. CD4⁺ T cells from patients also exhibited hypermethylation at TOLLIP and S100A6, genes involved in innate immune and inflammatory regulation. Collectively, these data demonstrate that CD4⁺ T cells from CP/CPPS patients display coordinated epigenetic remodeling across multiple immune-regulatory loci, including IL10 and FOXP3, suggesting that regulatory dysfunction may originate within this T cell compartment.

**Figure 3.**
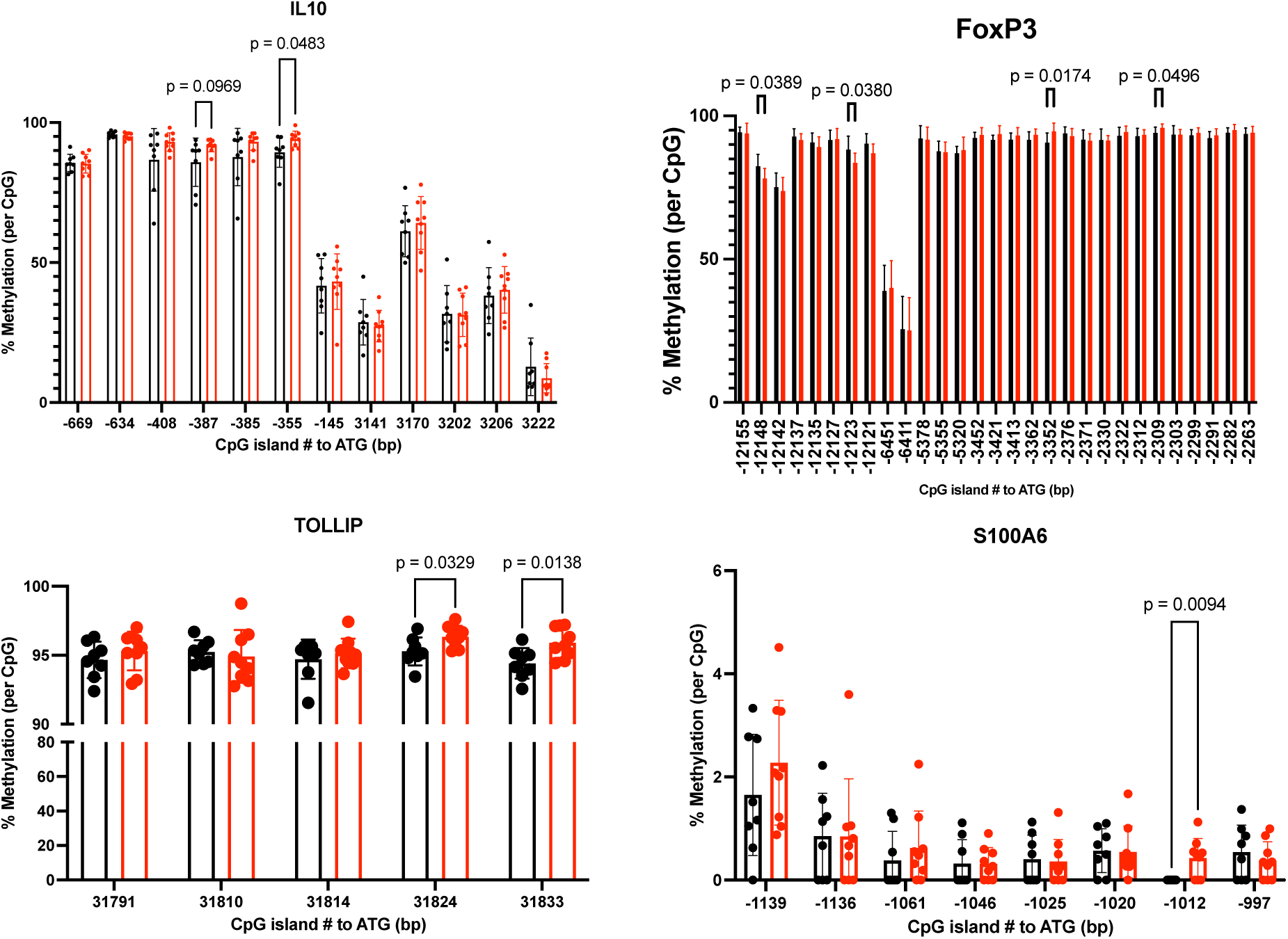
Expanded methylation changes in purified CD4⁺ T cells. DNA methylation analysis was performed on purified CD4+ T cells isolated from the peripheral blood of CP/CPPS patients (n = 9) and age-matched healthy controls (n = 8) using the EpigenDx Human FOXP3 array. Methylation at multiple CpG sites was assessed and compared between groups, with specific CpG islands identified relative to the ATG start site. Statistical comparisons were performed using two-tailed unpaired t-tests; individual p-values are shown.

### Functional consequences of IL10 hypermethylation

To determine whether IL10 expression is epigenetically regulated, PBMCs from healthy donors were treated with the DNA methyltransferase inhibitor azacitidine (AZA). AZA treatment produced a time-dependent increase in IL10 mRNA expression, confirming that IL10 is normally under epigenetic control through DNA methylation (Figure 4a). Because IL10 is typically induced by proinflammatory stimuli to dampen ongoing immune responses, we next assessed the functional impact of IL10 promoter hypermethylation observed in CPPS patients. PBMCs from healthy controls responded to lipopolysaccharide (LPS) stimulation by producing robust levels of IL10, whereas PBMCs from CPPS patients showed a markedly diminished IL10 response (Figure 4b). These findings suggest that IL10 promoter hypermethylation contributes to a functional impairment in the anti-inflammatory capacity of immune cells in CPPS.

**Figure 4.**
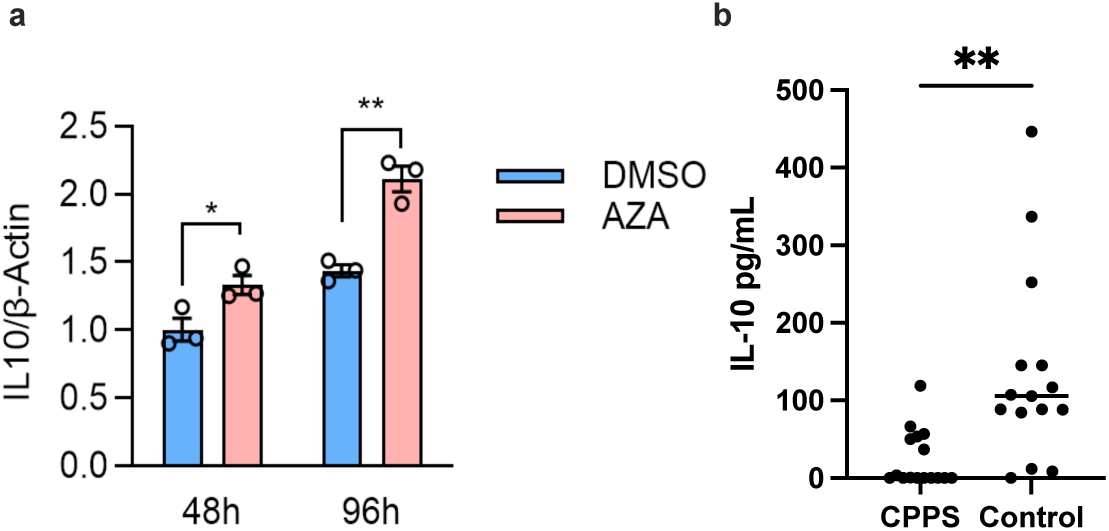
Functional consequences of IL10 hypermethylation. (a) PBMCs from CP/CPPS patients were treated with 1 μM azacitidine (AZA) for 48 or 96 hours to inhibit DNA methylation, followed by qRT-PCR for IL10 expression. (b) PBMCs from CP/CPPS patients were stimulated in vitro with LPS (1 μg/mL, 24 h), and IL-10 levels in supernatants were quantified by ELISA. Statistical comparisons were performed using two-tailed unpaired t-tests; *p<0.05, **p<0.01.

### *In vivo* validation of epigenetic therapy in the EAP model

To determine whether restoring regulatory function through demethylation could alleviate pelvic pain in vivo, we employed the experimental autoimmune prostatitis (EAP) model, a well-established murine model of CP/CPPS (37–42). Mice were immunized with prostate antigen and allowed to develop chronic pelvic pain, quantified by mechanical allodynia using von Frey testing. Administration of recombinant IL-10 beginning on day 22 of disease did not alter pain responses (Fig. 5a), indicating that cytokine supplementation alone was insufficient once regulatory defects were established. In contrast, treatment with the DNA methyltransferase inhibitor azacitidine (AZA) initiated on day 28 significantly reduced pelvic allodynia within one week of therapy (Fig. 5b). These data indicate that pharmacologic demethylation can ameliorate pain behaviors in the EAP model, consistent with restoration of regulatory pathways disrupted by DNA hypermethylation.

**Figure 5.**
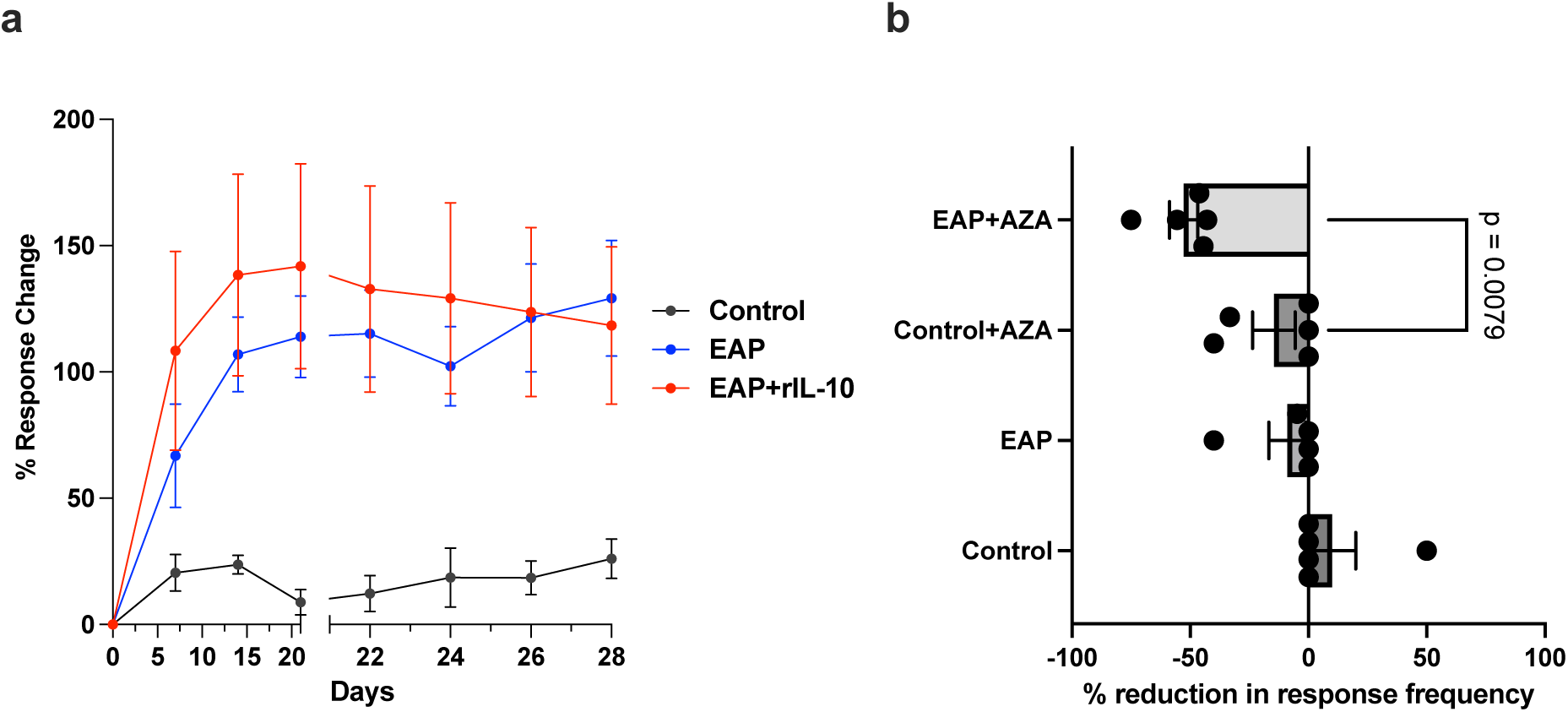
*In vivo* validation of epigenetic therapy in the EAP model. (a) In vivo therapeutic studies were conducted in experimental autoimmune prostatitis (EAP) mouse models. NOD/ShiLtJ mice (6–7 per group) were immunized with rat prostate antigen and treated with recombinant mouse IL-10 (1 μg/day, i.p., days 22–27). Mechanical allodynia was assessed by von Frey filament testing. (d) In a separate EAP cohort, C57BL/6J mice (5 per group) received 5-AZA (2.5 mg/kg/day, i.p., days 28–34), and mechanical allodynia was assessed before and after treatment. Statistical analyses were performed using ANOVA with multiple comparisons; individual p-values are shown.

## Discussion

Emerging evidence indicates that epigenetic mechanisms, particularly DNA methylation, contribute to CP/CPPS pathogenesis. Prior studies identified hypermethylation of estrogen receptor genes (ESR1/ESR2) linked to elevated estradiol levels [23], sperm DNA fragmentation and altered protamine ratios [2], and differential neuroinflammatory gene expression in CP/CPPS cohorts [32]. However, the functional significance of epigenetic changes within immune cells remains underexplored. Our study addresses this by identifying immune-specific methylation alterations in IL10 and FOXP3, linking them to immune dysregulation and chronic pain in CPPS.

Our findings show increased CD4⁺ T cells and elevated RORγT expression in prostatic urine, indicating Th17 skewing—consistent with prior studies implicating IL-17 in CPPS pathogenesis [20]. Concurrently, reduced FOXP3 expression suggests impaired Treg function. We identified hypermethylation at multiple CpG sites in the IL10 promoter, potentially silencing expression of this key anti-inflammatory cytokine. This is supported by prior work showing lower frequency of IL-10–producing genotypes in CPPS patients (30.6% vs. 12.1%, *p* = 0.007), which correlated with inflammation and treatment response [32].

Hypomethylation of the ITGAL promoter suggests increased leukocyte adhesion capacity, while altered methylation in PDL2, TNFα, and MYC indicates a broader inflammatory activation profile. Interestingly, no FOXP3 methylation change was detected in PBMCs; however, purified CD4⁺ T cells revealed hypermethylation at CNS1 and CNS2—regions critical for FOXP3 expression and Treg stability.

We also observed consistent methylation changes in TOLLIP and S100A6, genes linked to innate immunity and inflammation. These patterns suggest distinct immune cell subsets contribute to CPPS pathophysiology. Comparison of PBMCs and CD4⁺ T cells highlighted the importance of cell-type resolution, as several inflammatory gene methylation changes appeared specific to non-CD4⁺ populations.

Functionally, IL10 expression was restored in PBMCs following azacitidine (AZA) treatment, confirming its repression via DNA methylation. Moreover, IL10 induction following LPS stimulation was blunted in CPPS patients. In vivo, recombinant IL10 failed to reverse pelvic pain in a mouse model, whereas AZA significantly improved pain sensitivity, emphasizing the therapeutic potential of targeting methylation over cytokine supplementation.

These findings align with studies in other autoimmune diseases. IL10 promoter hypermethylation has been linked to reduced cytokine expression in a number of autoimmune diseases including SLE and RA [40] Similarly, FOXP3 hypermethylation at the TSDR impairs Treg function in RA and other immune and non-immune disorders [38; 39]. Our findings support the view that CPPS shares immune-epigenetic features with other chronic inflammatory disorders and may reflect systemic dysregulation rather than being limited to the pelvic region. Recent studies, including the MAPP Research Network, have emphasized the concept of “widespreadness,” showing that a subset of UCPPS patients experience multi-site pain and overlap with other chronic pain and autoimmune disorders such as fibromyalgia, irritable bowel syndrome, and chronic fatigue syndrome [9; 10; 14; 30]. The epigenetic alterations we identified—particularly in regulatory genes like IL10 and FOXP3—may define a subgroup of patients with broader immune dysfunction. Prior work has shown that such patients often have heightened systemic inflammation and an increased burden of extra-pelvic symptoms [12; 24]. Future research should assess whether these epigenetic signatures are enriched in patients with co-morbid autoimmune conditions (e.g., lupus, rheumatoid arthritis), which would clarify links between immune dysregulation, epigenetic control, and the widespread clinical manifestations seen in UCPPS [36]. These studies would address underlying mechanisms and clarify patient phenotypes based on mechanism of action (MOA) for future clinical trial designs in UCPPS [33].

In summary, IL10 and FOXP3 promoter methylation may define an immune-activated CPPS endotype characterized by impaired regulatory capacity and persistent inflammation. Epigenetic reprogramming represents a promising therapeutic strategy for restoring immune balance in CPPS.

## Data Availability

All data produced in the present study are available upon reasonable request to the authors

## ACKNOWLEDGMENTS

*Funding:* This work was supported by the National Institute of Diabetes and Digestive and Kidney (NIDDK) grants R01DK108127 and R01DK124460 to Praveen Thumbikat. The funders had no role in study design, data collection and analysis, decision to publish, or preparation of manuscript.

